# Evaluation of the role of home rapid antigen testing to determine isolation period after infection with SARS-CoV-2

**DOI:** 10.1101/2022.03.03.22271766

**Authors:** Lisa A. Cosimi, Christina Kelly, Samantha Esposito, Scott Seitz, Cole Sher-Jan, Jacquelyn Turcinovic, Kelley Friedman, Michael Nashed, Jesse Souweine, Nicholas Fitzgerald, Stacey Gabriel, John H. Connor, Deborah Hung

**Affiliations:** Brigham and Women’s Hospital, Division of Infectious Diseases, Boston, MA, USA 02115; National Emerging Infectious Diseases Laboratories, Boston, MA, USA 02118; Department of Microbiology, Boston University School of Medicine, Boston, MA, USA 02218; Bioinformatics Program, Boston University, Boston, MA, USA 02215; The Broad Institute of MIT and Harvard, Cambridge, MA, USA 02142; Department of Molecular Biology and Center for Computational and Integrative Biology, Massachusetts General Hospital, Boston, MA UA 02115

## Abstract

**Importance:** Recent CDC COVID-19 isolation guidance for non-immunocompromised individuals with asymptomatic or mild infection allows ending isolation after 5 days if asymptomatic or afebrile with improving symptoms. The role of rapid antigen testing in further characterizing the risk of viral transmission to others is unclear.

**Objective:** Understand rates of rapid antigen test (RAT) positivity after day 5 from a positive COVID-19 test and the relationship of this result to symptoms and viral culture.

**Design:** In this single center, observational cohort study, ambulatory individuals newly testing SARS-CoV-2 positive completed daily symptom logs, and RAT self-testing starting day 6 until negative. Anterior nasal and oral swabs were collected on a subset for viral culture.

**Main Outcomes and Measures:** Day 6 SARS-CoV-2 RAT result, symptoms and viral culture.

**Results:** 40 individuals enrolled between January 5 and February 11, 2022 with a mean age of 32 years (range 22 to 57). 23 (58%) were women and 17 (42%) men. All were vaccinated. 33 (83%) were symptomatic. Ten (25%) tested RAT negative on day 6. 61 of 90 (68%) RATs performed on asymptomatic individuals after day 5 were positive. Day 6 viral cultures were positive in 6 (35%) of 17 individuals. A negative RAT or being asymptomatic on day 6 were 100% and 78% predictive respectively for negative culture, while improving symptoms was 69% predictive. A positive RAT was 50% predictive of positive culture.

**Conclusion and Relevance:** RATs are suboptimal in predicting viral culture results on day 6. Use of routine RATs to guide end of COVID-19 isolation could result in significant numbers of culture negative, potentially non-infectious individuals undergoing prolonged isolation. However, a negative RAT was highly predictive of being culture negative. Complete absence of symptoms was inferior to a negative RAT in predicting a negative culture result, but performed better than improving symptoms. If a positive viral culture is a proxy for infectiousness, these data may help further refine a safer strategy for ending isolation.

## Introduction

More than two years after COVID-19 was declared a global pandemic, there is an ongoing need for policies both to ensure society can function normally and to protect those with ongoing risk of severe disease, in particular unvaccinated or immunocompromised individuals. Recent CDC COVID-19 isolation guidelines for non-immunocompromised individuals with asymptomatic or mild infection were changed to recommend ending isolation after 5 days for those who were asymptomatic throughout or, for those who were symptomatic, if now afebrile for 24 hours with improving symptoms. This period of isolation is to be followed by 5 days of wearing a tight-fitting mask when around others^1^. Support for this approach includes pre-omicron viral isolation and transmission studies suggesting the risk of COVID-19 transmission is highest in the one day prior and 3 days after symptom onset^2,3^ and that replicable viral recovery is uncommon in non-immunocompromised individuals after day 10^4–9^. There is currently little publicly available data regarding how long an individual remains infectious in the setting of high vaccination rates and the more highly transmissible Omicron variant. Additionally uncertainties include the role rapid antigen testing and pace of symptom resolution in determining the optimal duration of isolation.

Detection of SARS-CoV-2 nucleic acids by polymerase chain reaction (PCR) for prolonged periods after initial infection is common, including for Omicron. Importantly, these positive tests do not necessarily correspond to the ongoing presence of replication competent virus or to persistent infectiousness^10^.

However, modelling data has suggested up to a third of individuals may remain infectious after day 6^11^. Some have advocated using a rapid antigen testing as a surrogate marker for an individual’s infectiousness. RAT positivity in unvaccinated individuals correlates with viral culture positivity (positive predictive value (PPV), 90%), however, the role of rapid antigen testing in defining isolation periods has remained an open question^12–14^. Subsequent CDC guidance incorporates the use of rapid testing on day 6 or later, if feasible, recommending that if the RAT remains positive, isolation should continue through day 10.

Early data suggest that a high proportion of people will have a positive RAT after day 5. Specifically, two recent cross-sectional studies reported 43% and 54% RAT positivity in health care workers and in the community, respectively, after day 5. These results imply that the use of rapid antigen testing would significantly lengthen isolation periods for many people^15,16^. However, we lack systematic data in the Omicron era on the relationship between RATs and viral culture, and thus RATs and infectiousness after day 5, as they were not developed and approved for this use. While the full extent to which even viral culture truly reflects transmissibility remains uncertain, culture positivity has nevertheless played an essential role in guiding public health strategies in the midst of a pandemic. Thus, characterizing the relationship between RATs and viral culture during the recovery phase of COVID-19 infection is imperative for determining the utility of such tests in defining isolation periods.

### Objective

We sought to understand the performance of RATs after initial diagnosis by measuring the rates of RAT positivity after day 5. We correlated these findings with viral culture results in a healthy, vaccinated cohort with newly diagnosed COVID-19 to determine the potential role for rapid antigen testing after infection.

## Methods

From January 5 to February 11, 2022, we offered individuals testing positive for SARS-CoV-2 the opportunity to participate in a clinical research study evaluating the correlation of rapid antigen testing with culture positivity. The participants were drawn from a regularly scheduled twice-weekly program of COVID-19 testing done at the Broad Institute of MIT and Harvard. This testing was performed in the Broad’s CLIA-certified Clinical Research Processing Platform (CRSP) which has processed over 32 million COVID-19 tests since March of 2020. After release of the U.S. CDC’s most recent isolation guidance, individuals who tested positive and wished to return to campus prior to 10 days were supplied with at-home RATs. They were allowed to return if asymptomatic and/or afebrile for 24 hours with improving symptoms, and the RAT was negative on or after day 6 with day 0 being the date of their positive diagnostic test or first day of symptoms, whichever came earlier. To be eligible for this study, individuals needed to be an affiliate of the Broad, report a newly SARS CoV-2 positive test to the Broad and be 18 years or older. Beginning on day 6, enrolled individuals performed self-testing with the Flowflex™ lateral flow RAT - chosen because it is EUA approved for use in asymptomatic individuals^17^, uploaded a photo of the results and recorded daily presence or absence of symptoms (cough, fever, sore throat, difficulty breathing, chest tightness, fatigue, muscle aches, new loss of taste or smell, nausea, vomiting, diarrhea, runny nose, congestion, headache, other) until a negative test resulted. All individuals completed an online survey at entry to record recent COVID-19 contacts, presence and onset of symptoms, COVID-19 vaccination status, and dates of their most recent negative COVID-19 test. Cycle threshold values were available for individuals whose initial positive qRTPCR test was done at the Broad. Individuals followed standard institutional return-to-work protocol regardless of whether or not they participated.

A convenience sampling of observed, self-collected anterior nasal (AN) swabs and oral swabs were collected for 17 individuals on day 6 for viral culture (oral swabs collected as described by Marais et al)^18^. Swabs were immediately placed in viral transport media (Han Chang Medic™) and frozen at −20 °C until processing.

Viral culture was performed using Caco2 cells engineered to co-express Ace2 and TMPRSS2 which robustly support SARS-CoV-2 replication^19–21^. Caco2 cells were seeded into 24 well dishes in standard culture media (DMEM supplemented with 7% FBS, 50 ng/mL gentamycin and 0.25 ug/mL amphotericin B) overnight and grown to 60%-90% confluency. The following day 200 uL of viral transfer media from a patient sample was added to the culture media and allowed to incubate at 37 C. Three days after sample addition, cells were fixed in 10% formalin for 30 minutes at room temperature before removal from the BSL3. Cells were then permeabilized as previously described^22^ and analyzed for cytopathic effect (CPE) and evidence of viral replication by indirect immunofluorescence for the presence of SARS-CoV-2. Immunofluorescence was performed using an antibody that recognizes all strains tested of SARS-CoV-2 Nucleoprotein (Cell Signaling Technologies, E8R1L). Positive staining for SARS-CoV-2 growth was marked if cells showed strong immunolabeling in multiple cells and was considered negative if staining was not above mock infected cells. Using this approach, we have regularly cultured from AN and NP swabs with Ct values up to ∼32, including Omicron (500 genome equivalents; JHC unpublished results).

### Human Subjects

The research was reviewed and approved by the Mass General Brigham Institutional review board and by the Research Subjects Protection group at the Broad Institute. All enrolled individuals provided online written informed consent.

## Results

We enrolled 40 of 64 referred individuals newly diagnosed with COVID-19 between January 5 and February 11, 2022. The mean age was 32 years old (range 22 to 57), with 23 (58%) women and 17 (42%) men (Table 1). All individuals had completed a primary vaccine series and 36 (90%) had received a booster within a mean of 59 days prior to testing positive. The majority (33; 83%) reported symptoms, with an average of 1.2 days (range: −1 to 7) prior to testing positive. None were hospitalized. For the 29 whose positive test qRTPCR cycle thresholds were available because they were performed at the Broad Institute, mean and median Ct values were 26.5 and 28.9, respectively (Table 1). While these particular viral isolates were not sequenced, during the period of this study in the Boston area, 96-99% of sequenced isolates were the Omicron variant.

**Table 1.** (Cohort characteristics)

| **Table 1 (Cohort characteristics)** |  |  |  |
| --- | --- | --- | --- |
|  | **No. (%)** |  | **Range** |
| Total participants | 40 |  |  |
| Women | 23 (58) |  |  |
| Men | 17 (43) |  |  |
| Age (Mean, Range) | 34 |  | 22 to 57 |
| Race/ethnicity |  |  |  |
| white non-Hispanic | 21(53) |  |  |
| Black non-Hispanic | 3 (8) |  |  |
| Hispanic | 5 (13) |  |  |
| Asian | 7 (18) |  |  |
| Other/multiracial | 4 (11) |  |  |
| Known recent COVID-19 contact | 19 (48) |  |  |
| Have received primary COVID-19 vaccine series | 40 (100) |  |  |
| Have received COVID-19 vaccine booster | 36 (90) |  |  |
| Symptomatic at time of first positive test | 33 (83) |  |  |
|  | **Mean** | **Median** |  |
| Days since most recent vaccination dose | 59 | 54 | -2 to 184* |
| Days since most recent prior COVID-19 diagnostic test | 8 | 2 | 1 to 33 |
| Days of symptoms prior to testing positive | 1.2 | 1 | -1 to 7 |
| Ct value of positive COVID-19 PCR test (N =29) | 26.5 | 28.9 | 18.0 to 33.4 |

\*one individual received a booster 2 days after testing positive while awaiting result

Of the 40 participants, 10 (25%) tested negative on day 6 with a steady decline of daily positive results in the entire cohort out to 14 days, by which point all individuals tested negative. (Figure 1A). The median time to first negative RAT result was 9 days.

**Figure 1.**
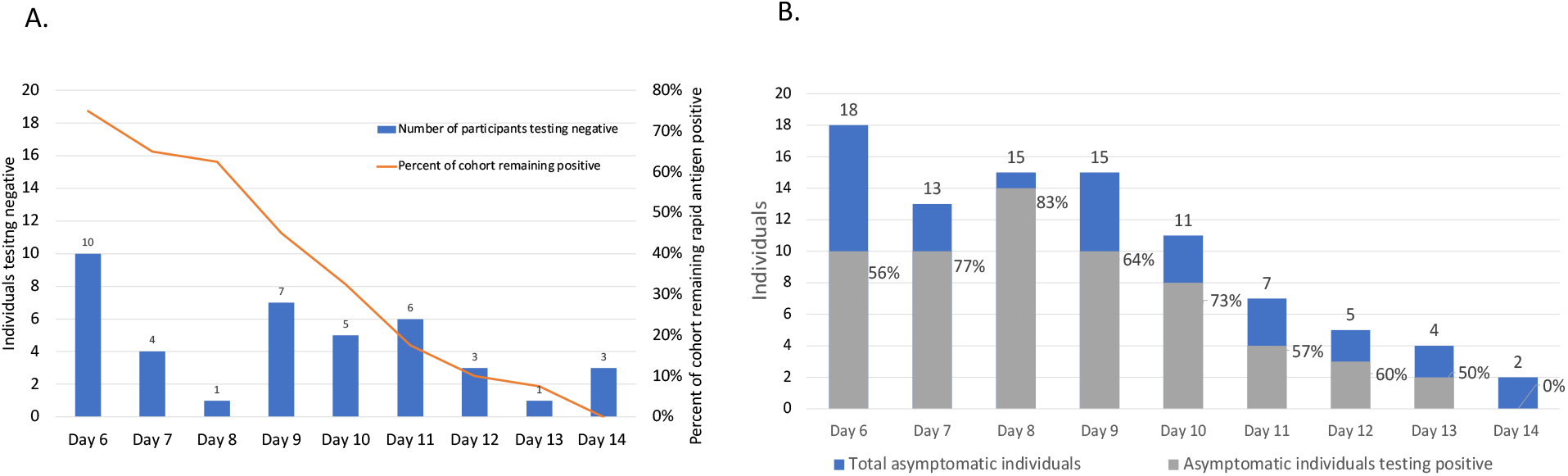
Days to first negative RAT for all participants and for asymptomatic individauls. **(A)** Daily count of individuals testing COVID-19 rapid antigen negative and proportion of cohort testing persistently positive (N=40). Individuals underwent daily home rapid antigen testing starting from day 6, after either their initial COVID-19 diagnosis or the start of symptoms, which ever came first. Shown in blue are the numbers of individuals who had their first negative on the designated day. In orange are the precent of remaining individuals still testing positive on each day. **(B)** Daily COVID-19 home RAT results in asymptomatic individuals. 161 total RATs were performed on 40 individuals starting on day 6 out to day 14. Among these, 90 tests were performed on individuals who were asymptomatic that day, with 61 being positive and 29 being negative (NPV 32%) (box top right). Shown in the graph for each day of testing are the numbers of asymptomatic individuals who tested negative (blue) and positive (grey). Percents indicate the percent of asymptomatic indviduvals testing positive on that day * 3 Individuals tested negative after day 10 (days 12, 13 and 14), but had missing testing data on one or more of the previous days. These individuals were left censored to negative at earliest possible negative day (day 11 (two individuals) and 12 (one individual)).

There was no correlation between time to first negative RAT and age, time since last vaccine, or Ct value at time of diagnosis. There was a trend toward earlier first negative test (8.1 vs. 9.2 days, *P*=0.14) for asymptomatic individuals, but this did not reach statistical significance (Supplemental Figure 1). Notably, the presence or absence of symptoms on the day of rapid antigen testing did not correlate with RAT results on that day (Figure 1B). Specifically, 10 of the 18 individuals who documented being asymptomatic on day 6 (56%), had a positive RAT. This same lack of correlation continued throughout the days of rapid testing, with 64-83% of asymptomatic individuals continuing to test positive through day 10. Out of 90 total RATs performed on asymptomatic individuals after day five, 61 (68%) were positive (Figure 1B).

To evaluate the relationship between a positive RAT and viral culture, we obtained AN and oral swabs for viral culture and qRTPCR testing for a subset of 17 individuals on day 6. Of these, 8 were asymptomatic on that day; 12 had a contemporaneous positive RAT while 5 had a negative test. Among the 17, 6 individuals (5 AN and 1 oral; 29%) were culture positive (Figure 2A).

**Figure 2.**
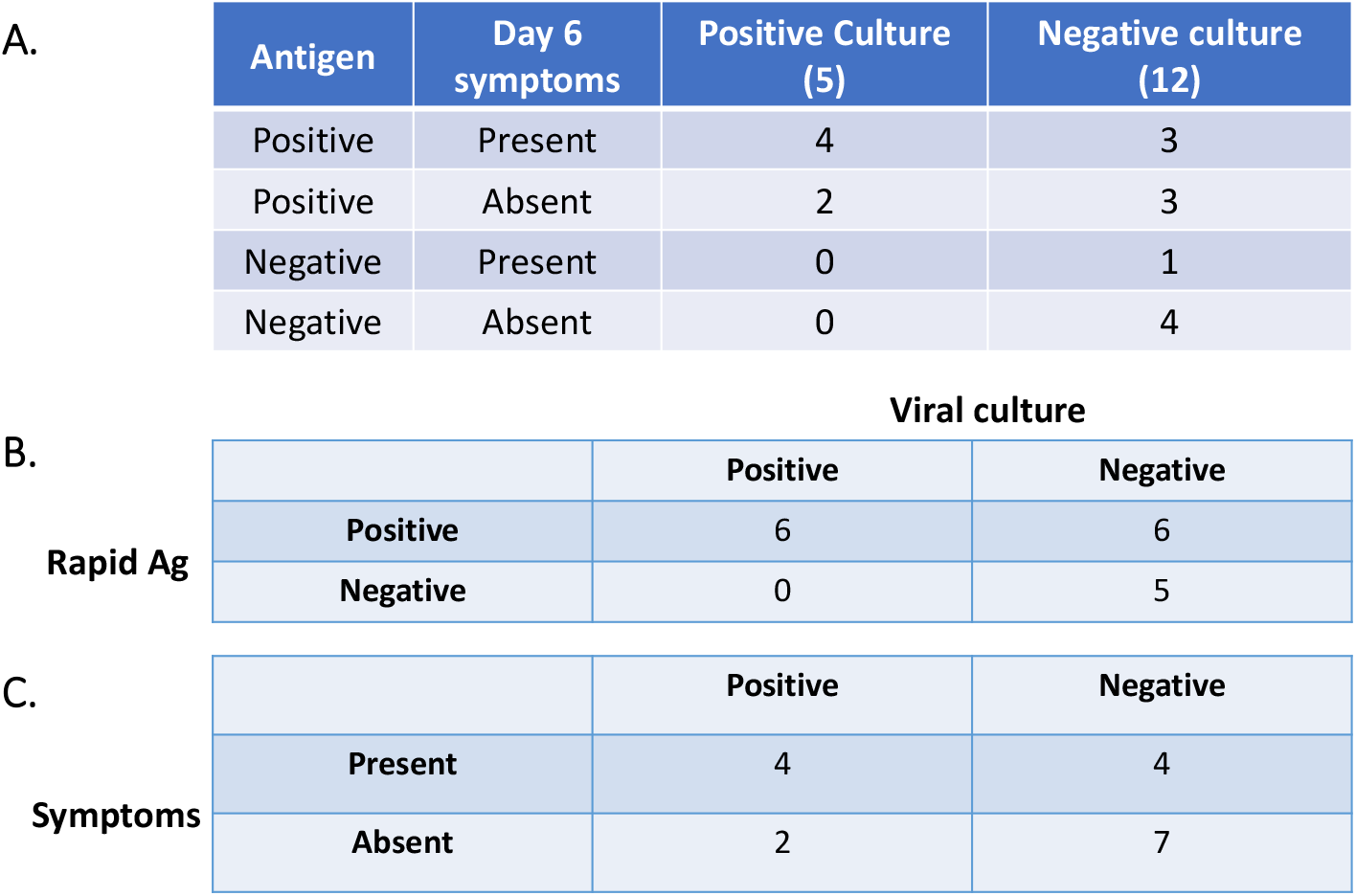
SARS-CoV-2 RAT result or symptoms as a predictor of viral culture. **(A)** Summary of individuals’ rapid antigen test results, symptoms, and viral culture results. Shown are numbers of individuals with a positive or negative viral culture as a function of their rapid antigen result and symptoms. **(B)** RAT result as a predictor of viral culture result. **(C)** Absence of symptoms on day 6 at the time of rapid antigen testing as a predictor of viral culture result.

All 6 (100%) of the culture positive samples were from individuals who had a positive RAT that day, and subsequently became RAT negative on days 7, 9, 10, 11, and 14 (2 individuals), respectively. Four (67%) reported improving (2) or unchanged (2) symptoms on day 6. Two were asymptomatic throughout (Figure 2A). However, we also observed 3 individuals who had both a positive RAT and were still symptomatic who had a negative culture. Thus, having both symptoms and a positive RAT had a PPV of 57% for being culture positive, while having a positive RAT or persistent symptoms individually had PPVs of 50%, respectively (Figure 2B, 2C). No asymptomatic individuals with a negative RAT were culture positive on day 6, resulting in a 100% NPV. Considered separately, no individuals with a negative RAT had a positive culture and two asymptomatics had a positive culture, resulting in 100% and 78% NPVs, respectively.

In total, 36 of the 40 enrolled and 13 of the 17 for whom we had culture data reported being asymptomatic or with decreasing (improving) symptoms and afebrile for 24 hours by day 6 (Figure 3A). By the CDC’s symptom-based guidelines, these individuals would have qualified for release from isolation, with masking. Compared to a requirement of the complete absence of symptoms for release, decreasing symptoms resulted in a lower NPV (69% vs 78%) and a similarly poor PPV (50%) (Figure 3B,C).

**Figure 3.**
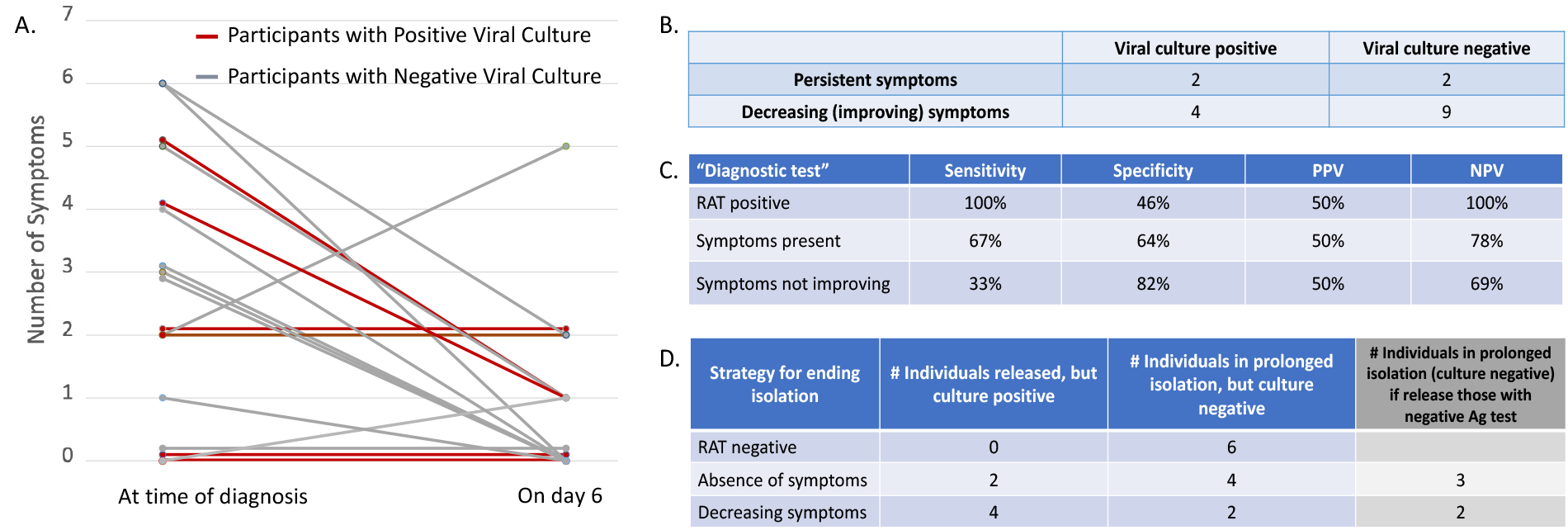
Decreasing (improving) symptoms as a predictor of viral culture result. **(A)** Numbers of symptoms reported by individuals at time of diagnosis and on day 6. Lines connect numbers of symptoms for each of the 17 individuals for whom we had culture data. Note, individuals who were asymptomatic both on day of diagnosis and day 6 were counted as part of the decreasing (improving) symptoms subgroup. **(B)** Decreasing or improving symptoms on day 6 at the time of rapid antigen testing as a predictor of viral culture result. **(C)** Summary of the performance of 3 approaches in predicting viral culture result – rapid antigen testing, presence of symptoms, and symptoms that are not improving. **(D)** The consequences for the 17 individuals for whom we had culture data, if each of the 3 strategies had been applied to determine when to end isolation, requiring either a negative RAT, the complete absence of symptoms, or declining (improving) symptoms on day 6. Consequences include the release of individuals who are culture positive and thus potentially infectious (false negatives) or the potentially unnecessarily prolonged isolation of some individuals who are culture negative (false positives). The false positive rate could theoretically be decreased by adding rapid antigen testing to guide release of persistently symptomatic individuals from isolation after day 5, if testing negative. Shown (grey column) is the impact that rapid antigen testing would have had in decreasing the numbers of false positives.

Based on the 17 individuals for whom we had culture data, a strategy for ending isolation based on a negative RAT would result in 6 individuals potentially being isolated longer than necessary, using a negative culture as a proxy for being “non-infectious”, but no release of any individuals who are culture positive (Figure 3D). Meanwhile, a strategy for ending isolation based purely on declining (improving) symptoms would result in 4 individuals being released who are still culture positive, while keeping 2 in isolation, potentially unnecessarily based on their negative culture. A strategy based on the complete absence of symptoms strikes a compromise between the other two strategies, resulting in the release of 2 culture positive individuals, at the expense of 4 individuals who are culture negative. Given its sensitivity (100%) and specificity (46%), a negative RAT could in theory reduce the numbers of individuals kept unnecessarily in prolonged isolation. In this cohort of 17 individuals, a negative RAT would have released one individual in the complete absence of symptoms strategy, reducing the number of potentially prolonged, isolated individuals to 3. (Figure 3D).

## Discussion

As isolation guidelines and masking mandates continue to evolve, it is critical to understand the risk of ongoing transmission in the early period post infection. Such information ideally can guide policies toward reducing transmission risk while also minimizing unnecessary isolation for people no longer infectious. In this study of mostly young, vaccinated individuals with mild COVID-19 and presumed Omicron variant, we found that most (75%) remain positive by rapid antigen testing on day 6, and a third through day 10. This is consistent with two recent cross-sectional studies in health care workers^15^ and in a community cohort of mixed-vaccinated individuals^16^. Similar to Lefferts et al, we saw a trend to earlier negative RAT in individuals asymptomatic at diagnosis that did not reach significance in this smaller, fully vaccinated cohort. Over one third in our cohort had initial symptoms that resolved by day 6, yet remained RAT positive on or after that day. Age, time from last vaccine dose, and the PCR Ct value at the time of diagnosis were not correlated with time to RAT negativity.

Culture positivity is believed to be the best proxy for infectiousness since only live virus can be transmitted. It has been proposed that detection of viral antigens might be an appropriate surrogate for viral culture, with previous reports correlating positive antigen testing with the ability to isolate virus. In this way rapid antigen testing contrasts with PCR, which can remain positive for a prolonged period even after viral cultures turn negative^12,14^. In this prospective study, we found that 35% of individuals still had culturable virus on day 6, which is consistent with previous modelling studies^11^. Looking at resolution of infection, we found a poor correlation between a positive RAT and positive viral culture (50% PPV), but excellent correlation (100% NPV) for a negative RAT. This is consistent with the interpretation that a positive RAT after day 6 and beyond does not always mean an individual is still infectious. Analogous to PCR testing, these positive antigen tests with negative cultures could reflect shedding of non-culturable, non-transmissible virus – albeit with a shorter duration of positivity compared to PCR as all individuals in our study were negative by RAT by day 14.

We acknowledge several limitations in this study, foremost is that it represents a relatively small cohort of young, vaccinated individuals with mild disease and presumed Omicron infection. These results would presumably vary in individuals who were older, unvaccinated or carried other co-morbidities, especially immunosuppression. The varying viral dynamics of the different variants would also inevitably affect the results of such a study. Additionally, the results are a function of the performance of self-collection and the technical assays used, including the specific RAT and its interpretation, and the method of culture. We intentionally used the Flowflex™ lateral flow RAT because it is FDA EUA approved for use in asymptomatic individuals. In addition, though RAT performance is in some studies less accurate in detection of the Omicron variant, the Flowflex™ has been shown to detect Omicron with reasonable sensitivity^23^. Similarly, while the decreased ability to culture Omicron has been reported^24^ our culture assay system utilizing Caco2 cells overexpressing Ace2 and TMPRSS2 has good sensitivity for culturing SARS-CoV-2 variants including Omicron,^19,21^ (JHC, unpublished data). Finally, and perhaps most significantly, both rapid antigen testing and culture are only proxies for transmission and correlates, not determinant, of infectiousness and transmissibility in vivo. While the degree to which viral culture completely and accurately reflects transmissibility remains an open question, the use of culture as the best current proxy has nevertheless been essential for rapidly learning about SARS-CoV-2 viral dynamics to inform public health strategies during the pandemic.

If culture positivity is indeed a good proxy for infectiousness, the drawback to a RAT-based strategy is that a potentially high proportion of individuals who test positive do not have a positive culture. This would practically mean unnecessarily prolonging isolation for these individuals. On the other hand, the strength of a RAT-based policy is that it releases no potentially infectious individuals. It also involves a more objective measurement compared to the subjective nature of symptom-based approaches. Meanwhile, as the vast majority of individuals have decreasing symptoms by day 6, an improving symptoms-based strategy, as is the centerpiece of current CDC guidelines, risks releasing culture positive, potentially infectious individuals prematurely, though the degree to which this is mitigated by mask-wearing for the subsequent 5 days is not known.

There are economic and social forces aiming to limit the period of isolation in order to return individuals to the workforce and to diminish the mental health consequences of isolation. These need to be balanced with the medical and public health motivation to protect those still at risk for contracting severe COVID-19. The important corollary is that the desired balance of these two forces and what infectious risk is considered tolerable are likely determined on an individual basis. The current CDC guidelines, which allow for stopping isolation provided symptoms are improving without requiring testing, are appropriate for settings which favor returning individuals to circulation in society; they allow for some potential risk of transmission. In our study, we found that complete abscence of symptoms, not simply improving symptoms, better predicted culture negativity, though not as well as a negative antigen test. For people with improving but residual symptoms, negative rapid antigen testing could provide additional reassurance about ending isolation. Selective RAT testing in these settings may strike a balance, between ending isolation of all individuals with improving symptoms, which could release potentially infectious individuals, and enacting a universal requirement of a negative RAT to end isolation, which could unduly extend isolation periods. Additional data is still needed from larger studies and from measurement of actual transmission. However, we hope these data provide initial insight into the potential strengths and limitations of rapid antigen testing as opposed to a purely symptom-driven strategy in guiding the end of isolation.

**Supplement Figure 1.**
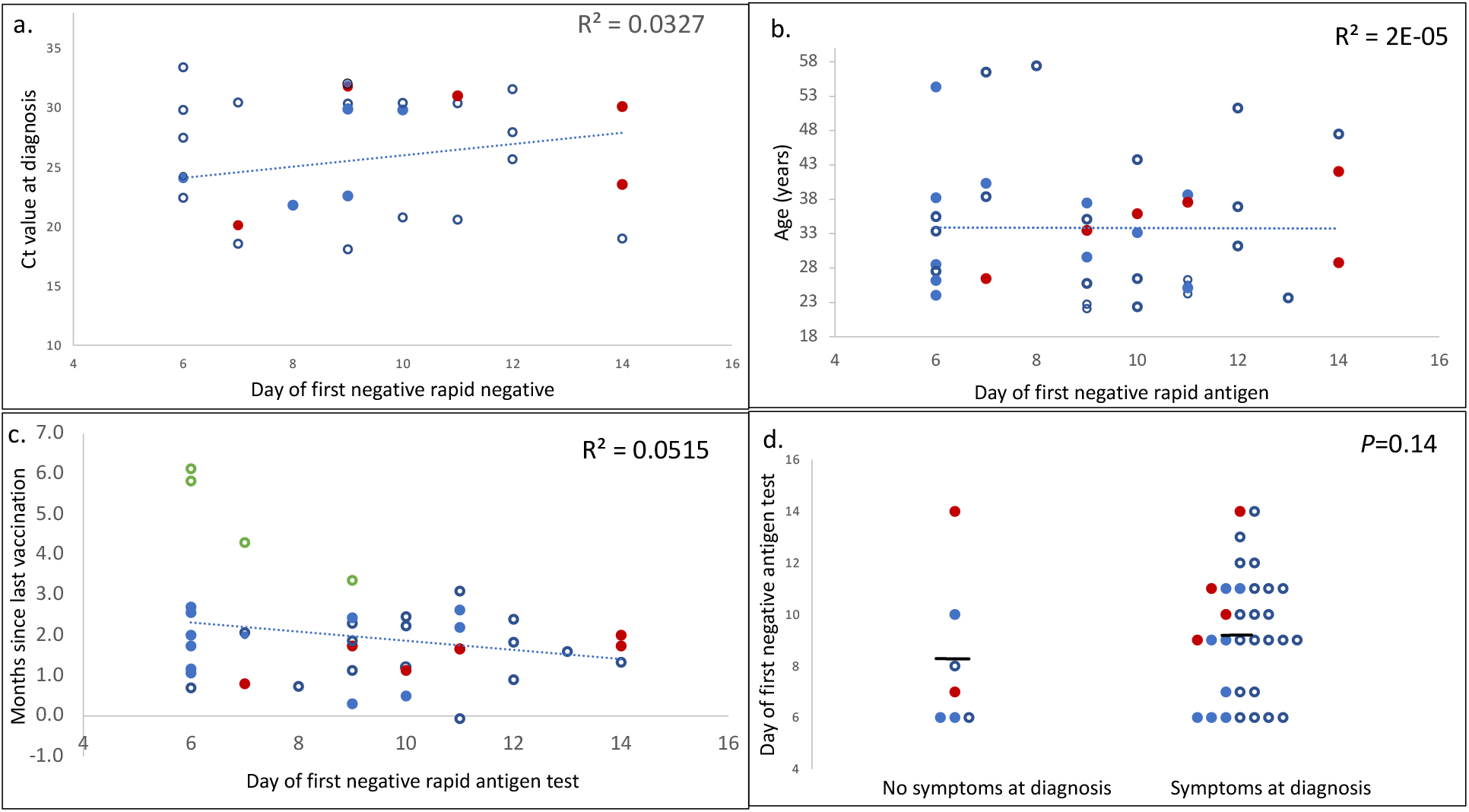
Relationship between days to first negative RAT and (a) Ct value at diagnosis, (b) participant age, (c) months since last vaccination, and (d) the presence/absence of symptoms at time of diagnosis. Each point represents a single individual in the study. Blue filled circles = individuals who had a sample taken for viral culture and were culture negative. Red filled circles = individuals who had a sample taken for viral culture and were culture positive. Blue open circles = all other individuals. In (c), the four individuals who had been vaccinated but not boosted are shown in green. *P*-value in (d) was calculated using the student-t test.

## Data Availability

All data produced in the present study are available upon reasonable request to the authors

## Author Contributions

Dr. Cosimi and Dr. Hung had full access to all of the data in the study and take responsibility for the integrity of the data and the accuracy of the data analysis.

Concept and design: Cosimi, Hung

Acquisition, analysis, or interpretation of data; all authors

Drafting of the manuscript: Cosimi, Hung, Connor, Esposito, Kelly

Critical revision of the manuscript for imortant intellectual content: Cosimi, Hung, Connor

Statistical analysis: Cosimi

Obtained funding: Hung, Souweine

Administrative, technical, or material support: all authors Supervision: Cosimi, Hung

Role of funder or sponsor in work: The Broad Institute’s Environmental Health and Safety Office informed individuals testing positive for COVID-19 of the opportunity to volunteer and consent for the study. The Procurement and Shipping Office mailed RATs to enrolled individuals.

## Conflict of Interest Disclosures

D.T.H. is a founder, consultant to, equity holder in, and inventor of technology licensed to Sherlock Biosciences and serves on the Scientific Advisory Board for Proof Diagnostics, both infectious disease diagnostic companies. Neither companies’ technologies were used in this work.

## Funding/Support

Costs were covered by operating funds of the Broad Institute of MIT and Harvard.

## Disclaimer

none

## Notes

### Author Declarations

The IRB of Mass General Brigham gave ethical approval for this work

## References

1. CDC, CDC. Quarantine & Isolation. Centers for Disease Control and Prevention. Published January 27, 2022. Accessed February 22, 2022. https://www.cdc.gov/coronavirus/2019-ncov/your-health/quarantine-isolation.html

2. He X, Lau EHY, Wu P, et al. Temporal dynamics in viral shedding and transmissibility of COVID-19. Nat Med. 2020;26(5):672–675. doi:10.1038/s41591-020-0869-5

3. Lauer SA, Grantz KH, Bi Q, et al. The Incubation Period of Coronavirus Disease 2019 (COVID-19) From Publicly Reported Confirmed Cases: Estimation and Application. Ann Intern Med. 2020;172(9):577–582. doi:10.7326/M20-0504

4. Wölfel R, Corman VM, Guggemos W, et al. Virological assessment of hospitalized patients with COVID-2019. Nature. 2020;581(7809):465–469. doi:10.1038/s41586-020-2196-x

5. van Kampen JJA, van de Vijver DAMC, Fraaij PLA, et al. Duration and key determinants of infectious virus shedding in hospitalized patients with coronavirus disease-2019 (COVID-19). Nat Commun. 2021;12(1):267. doi:10.1038/s41467-020-20568-4

6. Owusu D, Pomeroy MA, Lewis NM, et al. Persistent SARS-CoV-2 RNA Shedding Without Evidence of Infectiousness: A Cohort Study of Individuals With COVID-19. J Infect Dis. 2021;224(8):1362–1371. doi:10.1093/infdis/jiab107

7. Siedner MJ, Boucau J, Gilbert RF, et al. Duration of viral shedding and culture positivity with postvaccination SARS-CoV-2 delta variant infections. JCI Insight. 2022;7(2). doi:10.1172/jci.insight.155483

8. Bullard J, Dust K, Funk D, et al. Predicting Infectious Severe Acute Respiratory Syndrome Coronavirus 2 From Diagnostic Samples. Clin Infect Dis. 2020;71(10):2663–2666. doi:10.1093/cid/ciaa638

9. Arons MM, Hatfield KM, Reddy SC, et al. Presymptomatic SARS-CoV-2 Infections and Transmission in a Skilled Nursing Facility. N Engl J Med. 2020;382(22):2081–2090. doi:10.1056/NEJMoa2008457

10. Hay JA, Kissler SM, Fauver JR, et al. Viral dynamics and duration of PCR positivity of the SARS-CoV-2 Omicron variant. medRxiv. Published online January 1, 2022:2022.01.13.22269257. doi:10.1101/2022.01.13.22269257

11. Bays D, Whiteley T, Pindar M, et al. Mitigating isolation: The use of rapid antigen testing to reduce the impact of self-isolation periods. medRxiv. Published online January 1, 2021:2021.12.23.21268326. doi:10.1101/2021.12.23.21268326

12. Pekosz A, Parvu V, Li M, et al. Antigen-Based Testing but Not Real-Time Polymerase Chain Reaction Correlates With Severe Acute Respiratory Syndrome Coronavirus 2 Viral Culture. Clin Infect Dis. 2021;73(9):e2861–e2866. doi:10.1093/cid/ciaa1706

13. Korenkov M, Poopalasingam N, Madler M, et al. Evaluation of a Rapid Antigen Test To Detect SARS-CoV-2 Infection and Identify Potentially Infectious Individuals. Caliendo AM, ed. J Clin Microbiol. 2021;59(9). doi:10.1128/JCM.00896-21

14. Parvu V, Gary DS, Mann J, et al. Factors that Influence the Reported Sensitivity of Rapid Antigen Testing for SARS-CoV-2. Front Microbiol. 2021;12:714242. doi:10.3389/fmicb.2021.714242

15. Landon E, Bartlett AH, Marrs R, Guenette C, Weber SG, Mina MJ. High Rates of Rapid Antigen Test Positivity After 5 Days of Isolation for COVID-19. Infectious Diseases (except HIV/AIDS); 2022. doi:10.1101/2022.02.01.22269931

16. Lefferts B, Blake I, Bruden D, et al. Antigen Test Positivity After COVID-19 Isolation — Yukon-Kuskokwim Delta Region, Alaska, January–February 2022. MMWR Morb Mortal Wkly Rep. 2022;71(8):293–298. doi:10.15585/mmwr.mm7108a3

17. U.S. Food and Drug Administration. Flowflex COVID-19 Antigen Home Test. Published online February 18, 2022. https://www.fda.gov/media/152700/download

18. Marais G, Hsiao N yuan, Iranzadeh A, et al. Saliva swabs are the preferred sample for Omicron detection. medRxiv. Published online January 1, 2021:2021.12.22.21268246. doi:10.1101/2021.12.22.21268246

19. Chen DY, Khan N, Close BJ, et al. SARS-CoV-2 Desensitizes Host Cells to Interferon through Inhibition of the JAK-STAT Pathway. BioRxiv; 2020. doi:10.1101/2020.10.27.358259

20. Wilhelm A, Widera M, Grikscheit K, et al. Reduced Neutralization of SARS-CoV-2 Omicron Variant by Vaccine Sera and Monoclonal Antibodies. Infectious Diseases (except HIV/AIDS); 2021. doi:10.1101/2021.12.07.21267432

21. Widera M, Mühlemann B, Corman VM, et al. Surveillance of SARS-CoV-2 in Frankfurt am Main from October to December 2020 Reveals High Viral Diversity Including Spike Mutation N501Y in B.1.1.70 and B.1.1.7. Microorganisms. 2021;9(4):748. doi:10.3390/microorganisms9040748

22. Ruedas JB, Arnold CE, Palacios G, Connor JH. Growth-Adaptive Mutations in the Ebola Virus Makona Glycoprotein Alter Different Steps in the Virus Entry Pathway. Dutch RE, ed. J Virol. 2018;92(19). doi:10.1128/JVI.00820-18

23. Bekliz M, Adea K, Alvarez C, et al. Analytical sensitivity of seven SARS-CoV-2 antigen-detecting rapid tests for Omicron variant. medRxiv. Published online January 1, 2021:2021.12.18.21268018. doi:10.1101/2021.12.18.21268018

24. Fall A, Eldesouki RE, Sachithanandham J, et al. A Quick Displacement of the SARS-CoV-2 Variant Delta with Omicron: Unprecedented Spike in COVID-19 Cases Associated with Fewer Admissions and Comparable Upper Respiratory Viral Loads. Infectious Diseases (except HIV/AIDS); 2022. doi:10.1101/2022.01.26.22269927

